# Accuracy of diagnostic codes and algorithms used to identify connective tissue diseases in electronic health records and administrative databases: systematic review and meta-analysis

**DOI:** 10.64898/2026.09.23.26363795

**Authors:** Constanza Saka-Herrán, Thrshith Mani Prabu Kumar, Danielle Keane, Pierre Albert, Barbara Clyne, Caroline McCarthy, Gráinne Tynan, Nikki Dunne, Michelle Flood, Eoghan McCarthy, Frank Moriarty

## Abstract

**Objective:** To assess the accuracy of codes and algorithms used to identify selected connective tissue diseases (CTDs) in electronic health records (EHRs) and administrative databases.

**Methods:** We searched MEDLINE, Embase, and CENTRAL databases for studies that validated case definitions in EHRs against a reference standard, including rheumatologist-confirmed diagnosis or clinical classification criteria. Title/abstract screening, full-text review, data extraction, and quality appraisal were independently performed in duplicate. Findings were synthesised narratively and through a bivariate random-effects meta-analysis of sensitivity and specificity.

**Results:** A total of 38 studies were included. Systemic lupus erythematosus (SLE) was the most frequently studied disease, followed by systemic sclerosis (SSc). Across diseases, progressively restrictive algorithms generally improved positive predictive value (PPV), although the optimal strategy varied by disease. In SLE, multiple diagnostic codes provided the most favorable overall performance, whereas adding clinical data provided limited incremental benefit. In SSc, disease-specific clinical features, particularly Raynaud’s phenomenon, improved case identification, while specialist or inpatient-based algorithms performed better for polymyositis/dermatomyositis, although evidence was limited. For SLE, meta-analysis yielded pooled sensitivity of 0.90 (95% Cis 0.70-0.97), specificity of 0.83 (95% CIs 0.54-0.95), and PPV of 0.75 (95% CIs 0.59-0.86). Evidence for Sjögren’s Disease was limited to one study, and no studies evaluated MCTD/UCTD.

**Conclusions:** Optimal EHR case-identification strategies vary by disease and should be tailored to disease-specific clinical and diagnostic characteristics rather than algorithm complexity alone. Future research should develop and externally validate robust, standardised algorithms in representative populations, using rigorous validation methods, and assess the added value of longitudinal EHR data, biomarkers, and computational phenotyping approaches.

**Significance and innovations:**

- Current evidence supports disease-specific approaches to EHR-case identification: repeated diagnostic codes provided the most favorable performance for SLE, whereas disease-specific clinical features improved identification of SSc and specialist or inpatient-based definitions showed better performance for PM/DM, although evidence for the latter was limited to four studies.
- Increasing algorithm complexity did not consistently improve case-identification accuracy, highlighting the importance of selecting data elements that reflect the clinical phenotype and diagnostic characteristics of each disease.
- Important evidence gaps remain, particularly for Sjögren’s Disease and MCTD/UCTD, as well as for externally validated algorithms incorporating longitudinal clinical information and computational approaches.
- This systematic review extends previous evidence by providing the most comprehensive synthesis to date of validated EHR algorithms across selected CTDs, incorporating newer multicomponent and computational approaches and evaluating the incremental contribution of laboratory, and medication data that had been identified as important gaps in earlier reviews, although evidence for these more complex algorithms remains limited.

## 1. Introduction

Connective Tissue Diseases (CTDs), including Sjögren’s Disease (SjD), systemic lupus erythematosus (SLE), and systemic sclerosis (SSc), are autoimmune diseases characterised by overlapping clinical manifestations, and reduced quality of life^1–3^. Epidemiological data are essential to understanding their natural history, risk factors, and outcomes^4^. Electronic health records (EHRs) enable large-scale study of CTDs by capturing such data from large populations^5,6^.

Accurate identification of patients with CTDs in EHRs is essential for research and healthcare planning. However, overlapping clinical features make them difficult to distinguish using clinical data alone^1–3^. Coding systems, such as the International Classification of Diseases (ICD), may not capture the diagnostic complexity of CTDs. Reliance on International Classification of Primary Care codes can lead to misclassification due to non-specific codes, and variability in recording practices^7,8^.

EHR-based algorithms for CTD identification show variable accuracy. An SLE algorithm combining ICD-9 codes, positive antinuclear antibody (ANA) tests, and medication achieved a positive predictive value (PPV) of 95% but a sensitivity of only 40%^9^, highlighting limitations of CTD-identification algorithms.

Despite these challenges, the accuracy of codes and algorithms to identify CTDs across healthcare systems has not been comprehensively evaluated. The most recent systematic review on SLE identification was published by Moores et al in 2013 and highlighted the need for improved validation methods and incorporation of clinical data alongside ICD codes^10^. Additionally, no reviews were identified by the authors that comprehensively has assessed the identification of other CTDs.

Since then, advances in EHR-based case identification have led to the development of more complex algorithms incorporating additional clinical data for SLE and other CTDs^11–13^ , highlighting the need for a new systematic review. Such evidence can inform best practice for research and clinical application and improve reliability of CTD identification in EHR. Therefore, we aimed to evaluate the accuracy of codes and algorithms used to identify CTDs in EHRs and other administrative databases.

## 2. Methods

The study protocol was registered on PROSPERO (CRD420251056943) and is reported in line with the PRISMA-DTA statement^14^. We focused on selected CTDs from the ICD-10 M30-M36 classification: SLE, SjD, SSc, polymyositis/dermatomyositis (PM/DM), and mixed/undifferentiated connective tissue disease (MCTD/UCTD).

### 2.1. Eligibility criteria

We included studies evaluating the accuracy of codes or algorithms used to identify CTDs in EHRs or administrative databases against a reference standard, including clinician-documented diagnosis, clinician-led chart review, other form of clinician confirmed diagnosis, or application of standard classification criteria (e.g., ACR/EULAR). Self-reported diagnoses as the reference standard were excluded^15^. Studies had to clearly report algorithm components and describe methods for accuracy assessment. Studies not primarily designed as algorithm validation studies were eligible if they provided sufficient data to calculate either sensitivity, specificity, PPV, and/or NPV. We excluded case reports, editorials, commentaries, and narrative reviews. No restrictions were placed on language or publication date.

### 2.2. Information sources & Search strategy

We searched MEDLINE via PubMed, Embase, and the Cochrane Central Register of Controlled Trials (CENTRAL). The search strategy was adapted from those developed by the Mini-Sentinel project^10,16,17^ supplemented with additional relevant terms. We examined PubMed MeSH terms and reviewed the terms on abstracts and full-texts of primary studies included in a previous systematic review (Supplementary Table S1). The strategy was validated by confirming that it retrieved all studies in the review by Moores et al^10^. The MEDLINE strategy was adapted for other databases. Search strategies are provided in Supplementary Table S2. Searches were conducted on June 9^th^, 2025.

### 2.3. Study records

#### 2.3.1. Data management

Records were managed using Covidence. Screening questions were developed based on eligibility criteria and pilot-tested by two reviewers, who independently screened the same 50 abstracts for calibration. Questions were refined and re-tested as necessary.

#### 2.3.2. Study selection

Two reviewers independently screened titles and abstracts against eligibility criteria. Potentially eligible records underwent full-text review. Disagreements were resolved through consultation with a third reviewer, and reasons for exclusion were recorded.

#### 2.3.3. Data collection process and data items

Data were extracted using a standardised template based on STARD criteria^18^ adapted for administrative databases (availability of data statement). Extracted data included study characteristics, data source and population, CTD studied, algorithm components, reference standard, and reported accuracy measures. Full descriptions of data items are provided in Supplementary Material S3.

### 2.4. Diagnostic accuracy measures

The primary measures of diagnostic accuracy were sensitivity, specificity, PPV, and NPV. When these measures were not reported, they were calculated from the available two-by-two contingency tables. Accuracy estimates were classified as high (≥80%), moderate (60%-79%), or low (<60%)^19^. Details provided in Supplementary Material S3.

### 2.5. Risk of bias and applicability

Risk of bias and applicability were assessed using QUADAS-2 across four domains: patient selection, index test, reference standard, and flow/timing^20^. Domains were rated as low, unclear, or high risk. Overall study bias followed QUADAS-2 guidance, with any domain rated as high resulting in an overall high risk of bias^14^. Assessments were performed independently by two reviewers.

### 2.6. Synthesis of results

Results were synthesised separately for each CTD. Definitions and algorithms were classified according to the number and type of codes (e.g., diagnostic, prescriptions, laboratory test) following the framework of Shrestha et al^21^, with modifications:

- Less restrictive algorithms: required a single diagnostic code from an outpatient visit or unspecified source (e.g., single ICD code).
- Restrictive algorithms: required multiple codes or the inclusion of specific combinations of codes or data types (e.g., procedures, prescriptions, laboratory tests, or hospitalisation codes).

Restrictive algorithms were categorised into subcategories according to their expected accuracy, components, and frequency of use across studies. These were: multiple diagnostic codes, ≥1 ICD code by provider/specialist or healthcare setting, ≥1 ICD code plus clinical data (laboratory or medication data), multicomponent algorithms, and computational algorithms (full definitions are provided in Supplementary material S4).

### 2.7. Meta-Analysis

We used a bivariate random-effects meta-analysis based on a generalised linear mixed-effects model (GLMM) to jointly pool sensitivity and specificity, accounting for within-and between-study variability and their correlation^22–24^. PPV estimates were pooled using a random-effects meta-analysis of proportions with the *meta* package in R (*metaprop* function), stratified by algorithm and reference standard.

Forest plots presented individual and pooled estimates for PPV, sensitivity, and specificity with 95% confidence intervals (CIs) across algorithm subgroups. Between-study heterogeneity was evaluated using I^2^ statistics derived from the variance components of the random-effects models. All analyses were conducted in R version 4.4.3.

## 3. Results

### 3.1. Study selection

A total of 3,678 records were identified. Of these, 38 studies met the inclusion criteria (Figure 1). Reasons for exclusion for the 77 full-texts are detailed in Supplementary Table S5.

**Figure 1.**
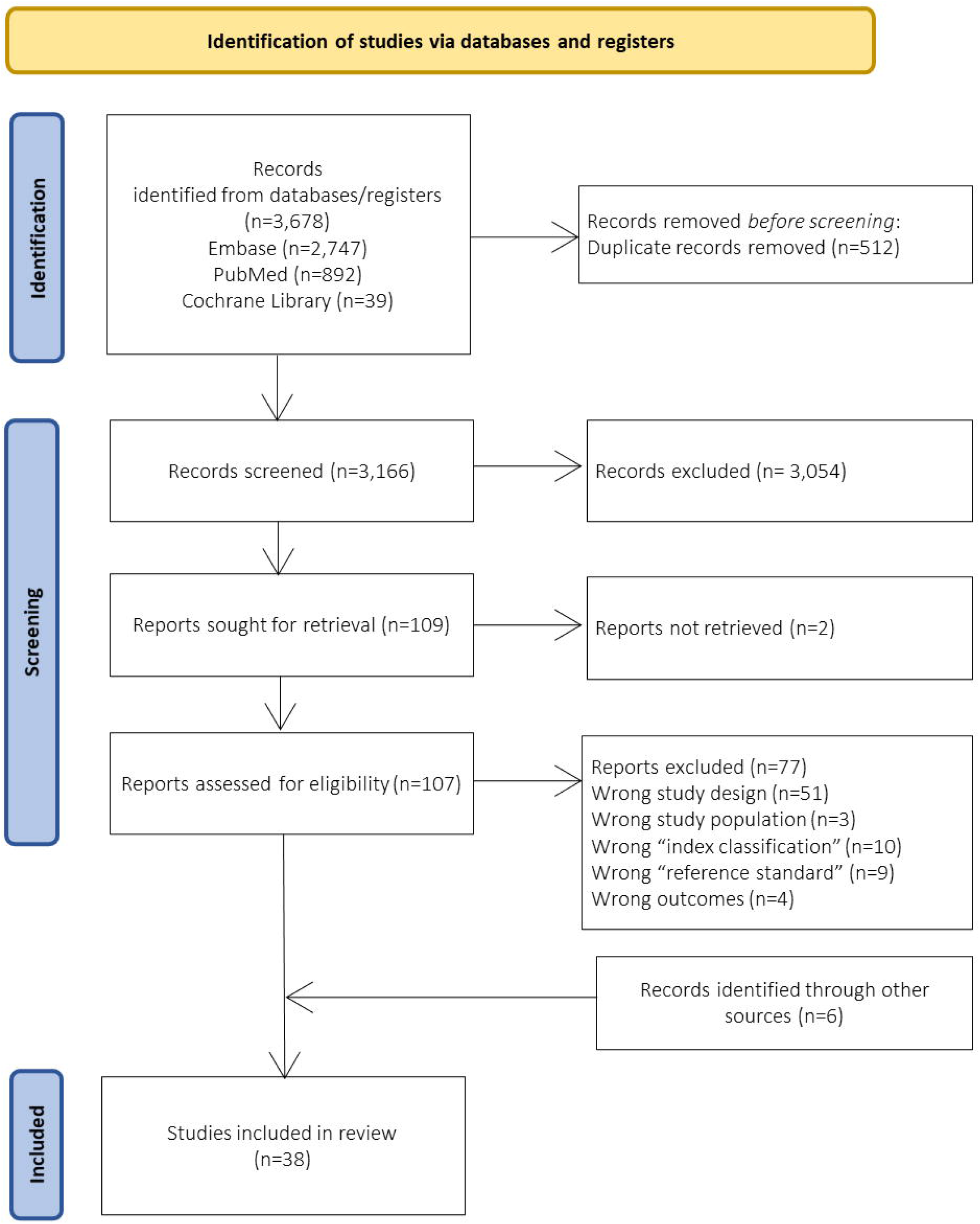
Flow diagram for study selection.

### 3.2. Study characteristics

Studies were published between 1997 and 2024, and conducted across North America, Europe, and Asia. The largest proportion originated from the USA (50%), followed by Northern Europe (26%). Most studies used a retrospective cohort design (71%), followed by cross-sectional (15.8%) and case-control (10.5%) approaches. Data sources included hospital-based EHRs^13,25–29^, national administrative registries^30,31^, insurance claims databases^32–36^, and population-based health data systems^37–40^ (Table 1 & Supplementary Table S6).

**Table 1.**
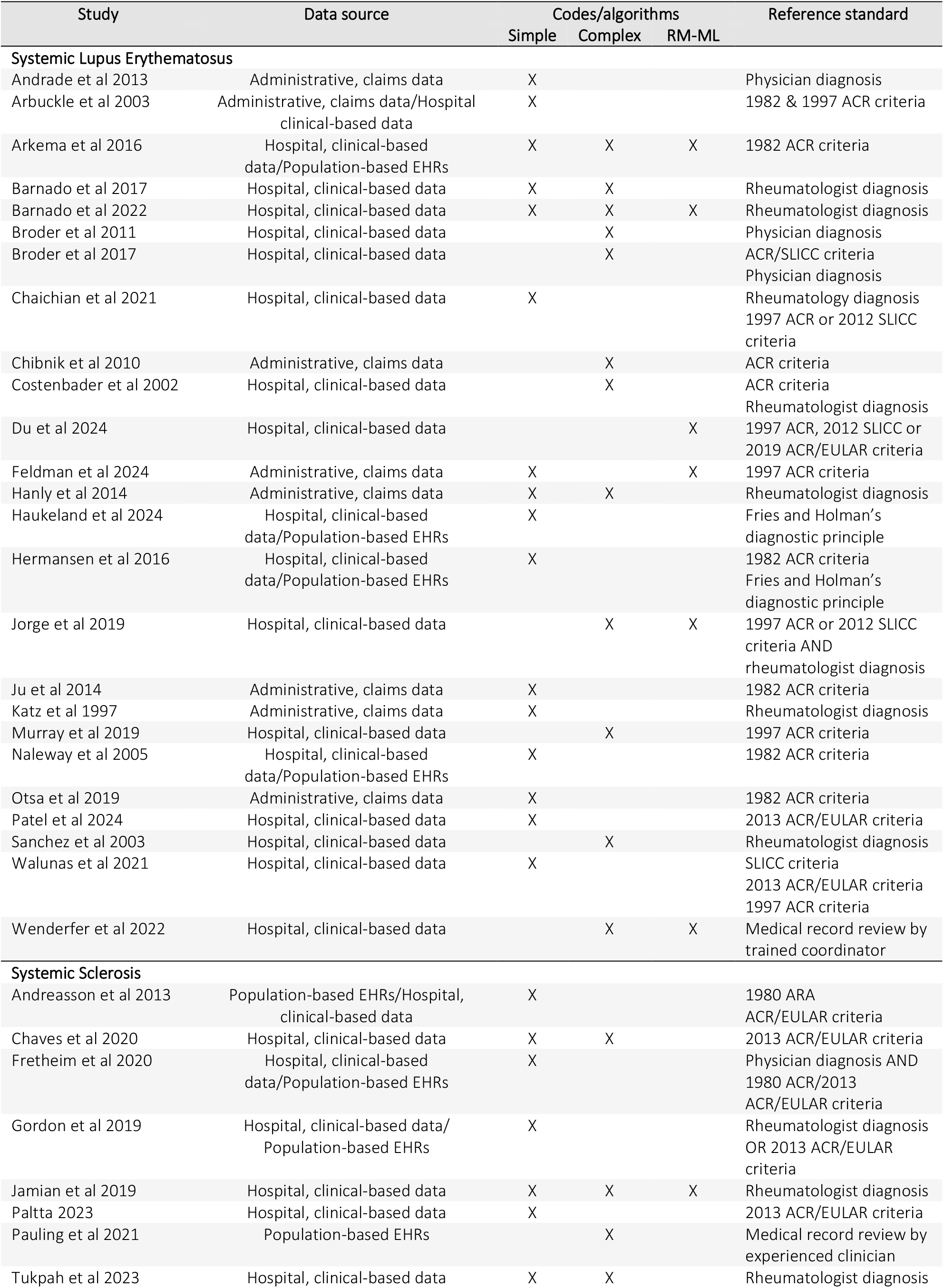
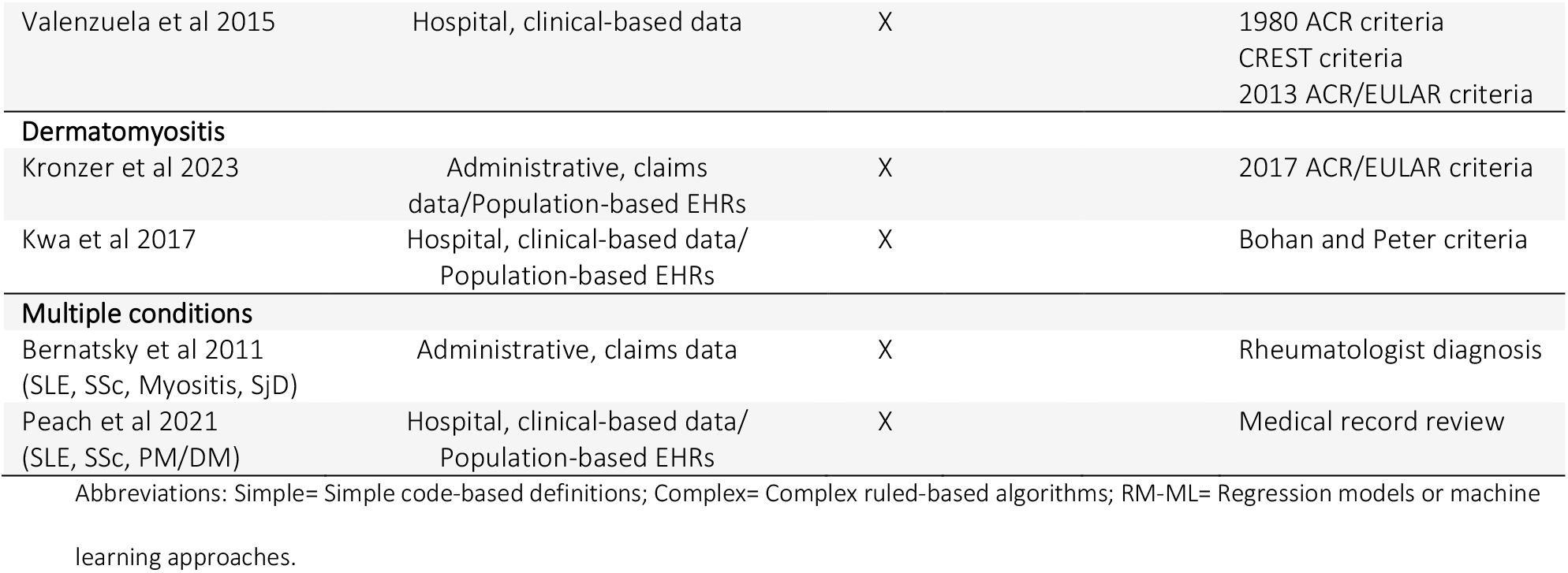
Summary of study characteristics across included studies (n=38)

SLE was the most frequently studied diagnosis (n=27), followed by SSc (n=11), myositis (n=4) and SjD (n=1). No studies evaluated MCTD or UCTD. Case identification was mainly based on ICD-9 (58%), and ICD-10 codes (53%), while a few relied on Read codes (3%)^37^, or SNOMED codes (3%)^39^; either alone or combined with additional structured data, including laboratory results (i.e., positive ANA, anti-ds-DNA) (26%), medication prescriptions (i.e., corticosteroids, disease modifying anti-rheumatic drugs) (16%), and other clinical attributes (e.g., renal failure, nephritis, Raynaud’s phenomenon) (29%).

Approximately half of studies used simple code-based definitions based on one or more diagnostic codes from administrative or EHR data^29,30,32,33,35,40–44^. Others applied rule-based algorithms combining diagnostic codes with laboratory values, medications, or other structured clinical data^9,11,13,36,38^ , while fewer used multivariable regression or machine-learning models, incorporating structured data and/or unstructured data^12,13,25,39,42,45^ (Table 1 & Supplementary Table S6).

Populations varied from clinical cohorts (<200 patients) to population-based samples (>10,000 individuals)^26,46^. Most included adults, while some focused on specific subgroups, including pediatric^39^, female^11,34^, hospitalised, or end-stage renal disease populations^41,47,48^. Several studies were limited to specialist rheumatology care or patients with multiple healthcare encounters, resulting in more selected populations with higher pre-test probability of disease^26,27,49^.

The most common reference standard for case validation was clinician medical record review, applying established clinical classification criteria such as ACR/EULAR (65.8%). Other studies relied on clinician documented diagnoses (42%), while a few applied either a rheumatologist-documented diagnosis or fulfilment of classification criteria as the reference standard^25,30,43^. Three studies relied on clinician or researcher medical record review without further specification for the criteria used^37,39,46^ (Table 1 & Supplementary Table S6).

### 3.3. Risk of bias and applicability concerns

The risk of bias varied across studies, with the greatest concerns identified in patient selection (high risk=21%, unclear risk=58%) and flow/timing domains (high risk= 31.5%, unclear risk= 55%). The reference standard domain was generally assessed as low risk of bias (low risk= 79%) (Table 2). Selection bias and concerns regarding applicability were greatest in studies using specialised, tertiary-care, or otherwise selected clinical populations. The most common source of high bias risk was flow/timing, due to partial verification of index-test positive cases and limited assessment of individuals without diagnostic codes, restricting evaluation of false negatives. The index test domain was generally rated as low (47%) or unclear risk (53%), due to incomplete methodological reporting (e.g., required number of code occurrences, the clinical setting of code assignment). A detailed domain-level assessment is provided in Supplementary Material S7.

**Table 2.**
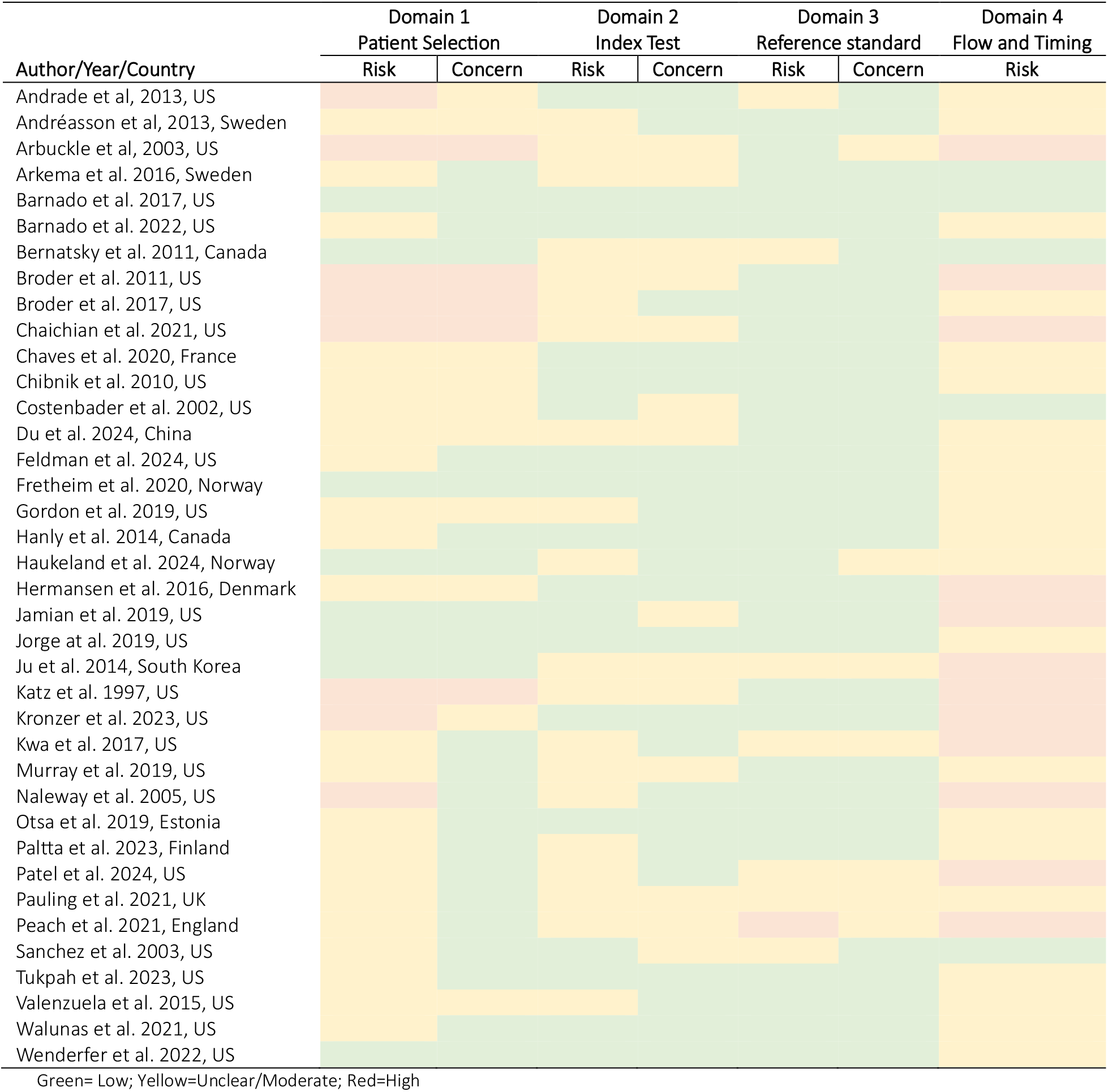
Risk of bias and applicability concerns across included studies (n=38)

### 3.4. Results of individual studies

Algorithms and results of individual studies (n=38) are detailed in Supplementary Material S8.

#### 3.4.1. Systemic Lupus Erythematosus (n=27)

The algorithms used to identify SLE varied widely across studies and could be broadly classified into six categories according to their level of restriction and data elements incorporated (Figure 2) (Full descriptions in Supplementary Material S4). Numerous studies (n=13) evaluated single diagnostic code algorithms, typically defined as at least one ICD code in outpatient or unspecified settings^32,33,35,44,49,50^. These definitions demonstrated high sensitivity, but their PPV was highly variable. For example, PPVs ranged from 9.5% in a large EHR-based cohort^49^ to approximately 67% in a nationwide administrative dataset^32^.

**Figure 2.**
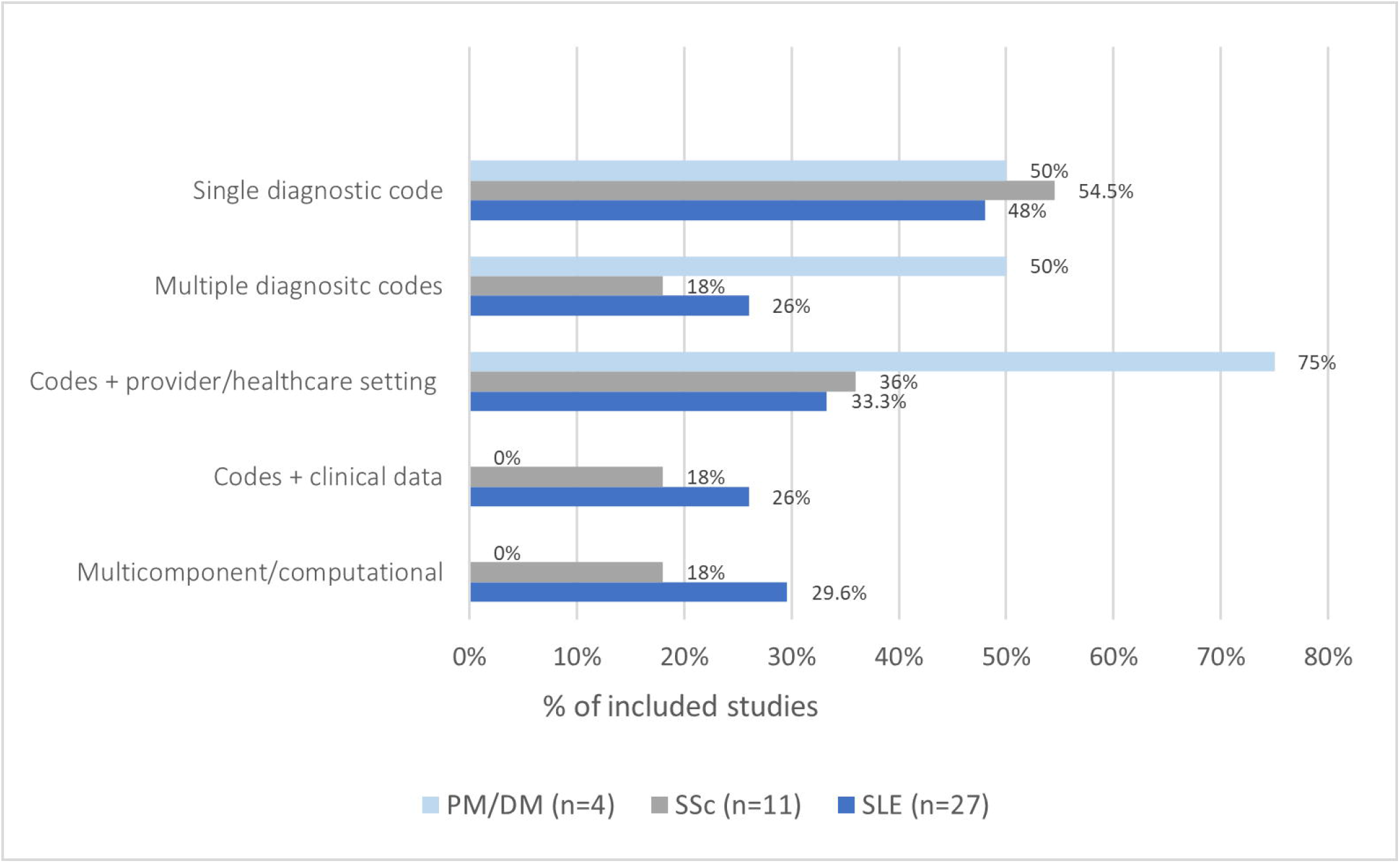
Frequency of algorithm categories across studies used to identify CTDs in electronic health records and administrative databases *(As individual studies frequently evaluated multiple algorithmic approaches, categories are not mutually exclusive)*.

Seven studies assessed algorithms requiring multiple diagnostic codes, defined as two or more codes recorded over time. Increasing the number of required codes (e.g., three or four) consistently improved specificity and PPV while reducing sensitivity^11,36,42,51^ .

Nine studies incorporated specialty information or healthcare setting to improve diagnostic accuracy. Algorithms restricting case identification to hospitalisations showed very high specificity and PPV but markedly lower sensitivity (e.g., specificity= 99.9%, PPV= 99.4%, sensitivity= 41% in Hanly et al 2014)^36^. Studies requiring diagnostic codes assigned by specialists, particularly rheumatologists, substantially improved case accuracy compared with codes recorded by non-specialists. These algorithms generally achieved high PPV (≥80%) without a substantial decrease in sensitivity (70-80%)^11,36^.

Several studies (n=7) evaluated algorithms combining diagnostic codes with laboratory values or medication data. These approaches frequently incorporated serologic markers (e.g., ANA/dsDNA) or medication treatment information. Algorithms combining a single ICD code with laboratory or medication data generally demonstrated moderate diagnostic accuracy (PPV <70%), indicating that the addition of clinical information alone was insufficient to substantially improve case identification. In contrast, algorithms requiring multiple diagnostic codes consistently achieved higher PPV and specificity, and the subsequent incorporation of laboratory or medication data provided only limited improvement^9,11,25^.

Eight studies developed multi-component rule-based algorithms and computational phenotypes, integrating multiple data elements simultaneously. These approaches ranged from structured clinical scoring systems to data-driven models. Examples include the use of the Boston Weighted Criteria in combination with diagnostic codes^52,53^, which demonstrated high sensitivity (>90%) and PPV (>80%) but moderate specificity (60-70%), reflecting emphasis on broader case capture. Rule-based approaches incorporated composite algorithms combining diagnostic codes with laboratory results, medication data, healthcare setting, specialist diagnosis, or clinical features, generally achieving high PPV (>85%)^9,45,54^.

Advanced computational phenotyping approaches using regression and machine-learning methods integrated structured and unstructured EHR data and generally demonstrated higher PPV and specificity than simpler code-based algorithms, although sensitivity varied across model approach^42^. These approaches were largely evaluated using internal validation or single-centre datasets, limiting their generalisability^12,25,26,42^.

#### 3.4.2. Systemic Sclerosis (n=11)

Algorithms used to identify SSc demonstrated consistent performance patterns according to algorithm restrictiveness (Figure 2). Single diagnostic code definitions (n=6 studies) showed variable PPV, ranging from 52% to 76%, depending on the reference standard applied, with higher estimates observed when validated against inclusive contemporary classification criteria, e.g., 2013 ACR/EULAR criteria^13,29,30,43,55^. Sensitivity was reported only by Jamian et al^13^, where a single diagnostic code achieved 98% sensitivity but only 52% PPV.

Algorithms requiring multiple diagnostic codes (n=2 studies) consistently improved accuracy. Jamian et al^13^, demonstrated that increasing the number of required ICD codes improved PPV from 52% for a single code to 91% for four codes, while sensitivity decreased modestly from 98% to 91%. Similar improvements were observed in Tukpah et al^28^, where requiring at least two ICD codes achieved a PPV of 78%.

Restricting case identification according to provider specialty or healthcare setting (n=4 studies) further improved performance. Algorithms requiring diagnosis by a specialist demonstrated higher PPV compared to less restrictive code definitions, as observed in Paltta et al (78%)^55^ and Andreasson et al (74%)^56^. Inpatient-based algorithms showed similar accuracy, with Peach et al^46^ reporting a PPV of 86.8% for hospitalised cases.

Only Jamian et al^13^ evaluated algorithms combining diagnostic codes with clinical and laboratory features, including ANA positivity, and documentation of Raynaud’s phenomenon (RP). Incorporation of these features improved diagnostic accuracy, with progressively more restrictive algorithms achieving higher PPV at the expense of sensitivity. The highest performing algorithm (≥3 ICD-10 codes and ANA positivity) achieved a PPV of 100% but a sensitivity of 50%.

Computational approaches (n=2 studies) further improved SSc case identification by integrating diagnostic code frequency with clinical information. Jamian et al^13^ used different approaches incorporating ICD code counts with clinical features, particularly RP and ANA positivity, with the random forest model achieving 84% PPV and 92% sensitivity, while identifying RP keyword as the strongest predictor of SSc. Similarly, Tukpah et al^28^ developed and internally validated text-processing algorithms combining ICD-10 codes with RP and esophageal involvement keywords. Adding RP keywords to a baseline definition of ≥2 ICD-10 codes increased PPV from 78% to 84%, with addition of esophageal involvement keywords adding no benefit (PPV=84%, sensitivity= 58%, specificity= 70%). Performance declined in the validation cohort (PPV= 76%).

Pauling et al^37^ developed a hierarchical case ascertainment strategy using UK primary care records, showing that Read codes alone overestimated SSc case frequency (PPV= 56.3%). Incorporating supporting clinical evidence, progressively increased PPV, reaching 85% in patients with the highest secondary evidence scores.

#### 3.4.3. Polymyositis/Dermatomyositis (n=4)

The accuracy of PM/DM identification algorithms varied with algorithm restrictiveness, although most studies reported PPV only (Figure 2). Single diagnostic codes (n=2 studies) showed limited and variable accuracy (range 5.6%-44%)^40,57^. Requiring multiple diagnostic codes (n=2 studies) modestly improved accuracy, with PPV increasing from 44% to 49% in Kronzer et al^40^, while Kwa et al^57^ reported 35% PPV for repeated outpatient codes, maintaining sensitivity (89%).

Restricting PM/DM identification according to specialty or healthcare setting (n=3 studies) achieved the highest improvements in diagnostic performance. Algorithms requiring diagnosis by rheumatologist or dermatologist improved PPV to 41%, preserving sensitivity (83%)^57^. Restricting identification to a principal diagnosis in inpatient discharges increased PPV to 95%, with a substantial decrease in sensitivity (23%). Similarly, Peach et al^46^ reported a PPV of 95% when restricting DM case ascertainment to inpatient-based codes and a PPV of 87% for identifying PM cases in hospital settings. This pattern was supported by the composite algorithm evaluated by Bernatsky et al^38^, which combined hospitalisation codes, repeated physician billing codes, and rheumatologist-assigned diagnosis to improve case identification validity.

#### 3.4.4. Sjögren’s disease (n=1)

One study evaluated the accuracy of an algorithm for identifying SjD in EHRs. Bernatsky et al^38^ assessed a composite administrative case definition based on hospitalisation diagnostic codes, ≥2 physician billing codes, or rheumatologist-assigned diagnostic code, demonstrating high diagnostic accuracy, with a sensitivity of 95.5%, specificity of 95.8%, and PPV of 73%.

### 3.5. Meta-Analysis

#### 3.5.1. Systemic Lupus Erythematosus

Using rheumatologist diagnosis of SLE as the reference standard, PPV varied substantially according to the case identification algorithm. The pooled PPV was 0.68 (95% CI 0.43-0.85) for a single ICD code, 0.75 (95% CI 0.59-0.86) for multiple ICD codes, 0.96 (95% CI 0.78-0.99) for inpatient ICD codes, 0.56 (95% CI 0.51-0.62) for ICD codes combined with laboratory data, and 0.59 (95% CI 0.48-0.69) for ICD codes combined with medication data. Substantial between-study heterogeneity was observed (I^2^ ranging from 69.5% to 92.2%), whereas no heterogeneity was detected for algorithms incorporating laboratory values (I^2^= 0%) (Figure 3A).

**Figure 3.**
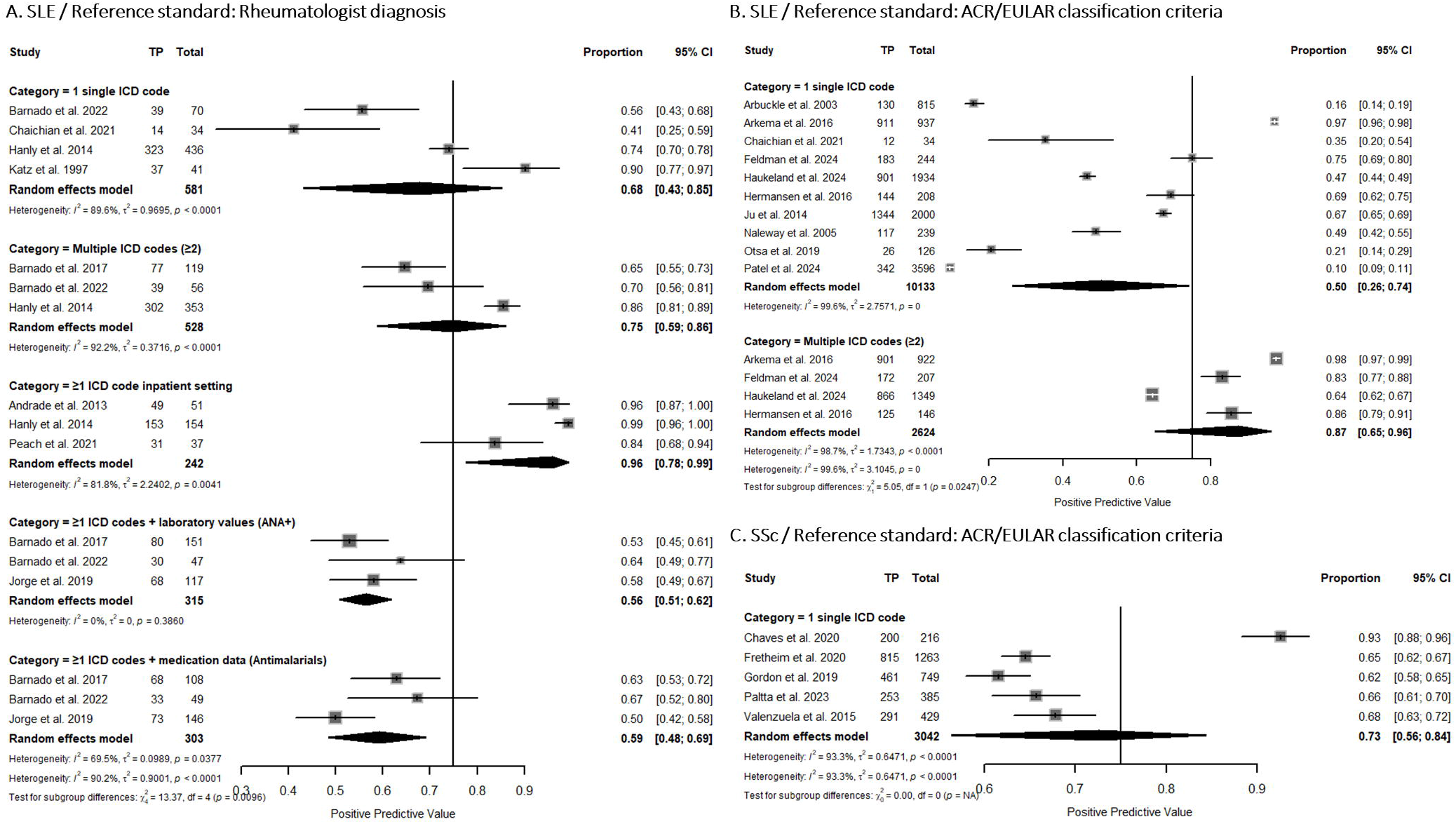
**A**. Forest Plot: Individual and pooled estimates of PPV across algorithms categories for SLE-case identification in EHRs using rheumatologist diagnosis as reference standard / **B**. Forest Plot: Individual and pooled estimates of PPV across algorithms categories for SLE-case identification in EHRs using ACR/EULAR classification criteria as reference standard / **C**. Forest Plot: Individual and pooled estimates of PPV across algorithm category for SSc-case identification in EHRs using ACR/EULAR classification criteria as reference standard.

Using ACR/EULAR classification criteria as the reference standard, algorithms based on multiple ICD codes demonstrated a higher pooled PPV (0.87, 95% CI 0.69-0.96) than algorithms relying on a single ICD code (0.50, 95% CI 0.26-0.74). Substantial heterogeneity was observed in both categories (I^2^= 99.6% and 98.7%, respectively). Meta-analysis for the remaining categories could not be performed due to the insufficient number of studies available (Figure 3B).

There were sufficient numbers of studies for bivariate random-effects meta-analysis of three algorithm categories using rheumatologist diagnosis as the reference standard. Algorithms based on multiple ICD codes demonstrated the highest pooled sensitivity, with an estimate of 0.90 (95% CI 0.70-0.97), and a pooled specificity of 0.83 (95% CI 0.54-0.95). Algorithms combining ICD codes with laboratory values yielded a pooled sensitivity of 0.83 (95% CI 0.74-0.90) and a pooled specificity of 0.63 (95% CI 0.40-0.81). Similarly, algorithms combining ICD codes with medication data showed a pooled sensitivity of 0.82 (95% CI 0.75-0.87) and a pooled specificity of 0.67 (95% CI 0.62-0.71) (Figure 4).

**Figure 4.**
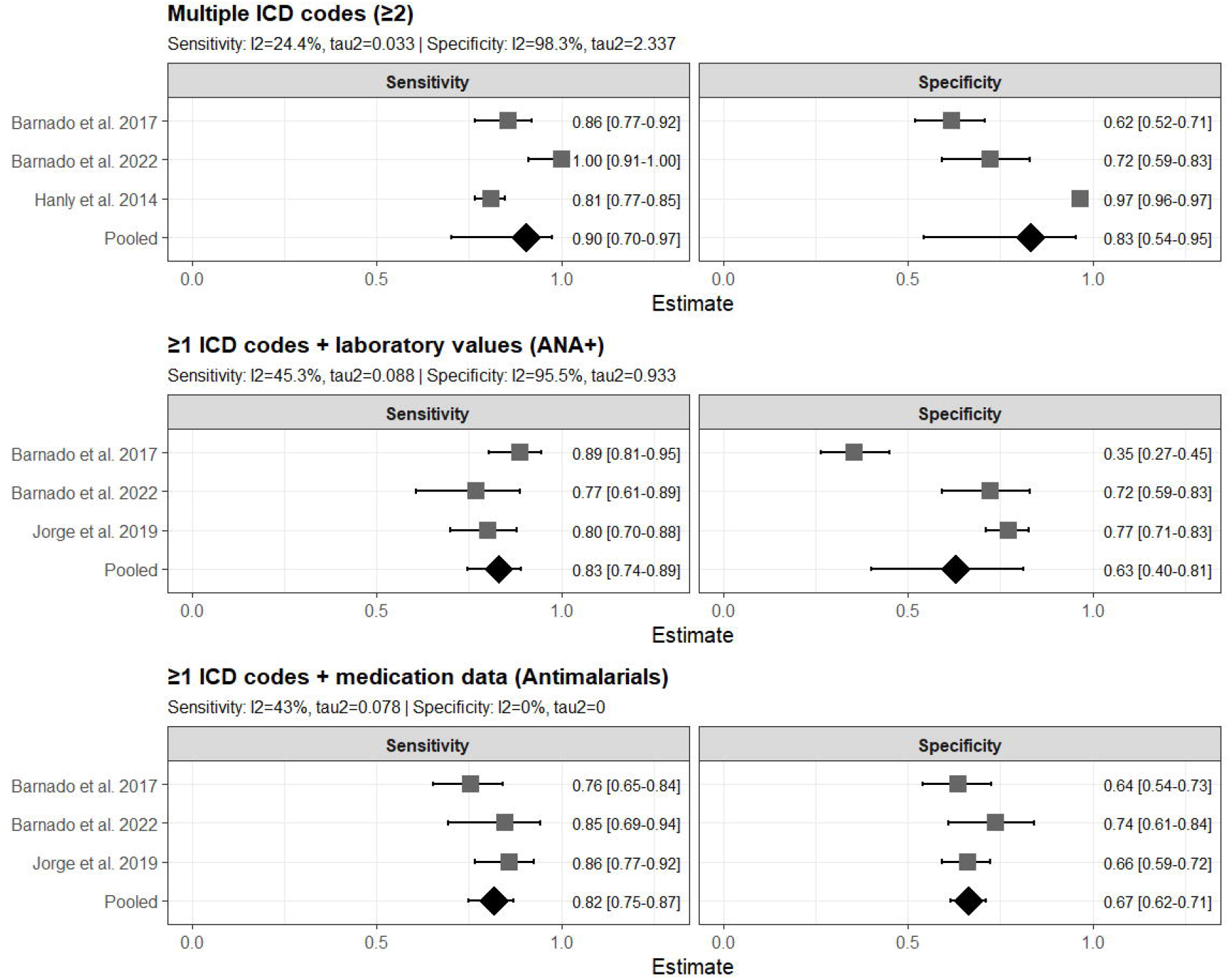
Forest Plot: Individual and pooled estimates of sensitivity and specificity across algorithms categories for SLE-case identification in EHRs using rheumatologist diagnosis as reference standard.

Between-study heterogeneity for sensitivity was low to moderate (I^2^ ranging from 24.4% to 45.3%). In contrast, substantial heterogeneity was observed for specificity among algorithms based on multiple ICD codes (I^2^= 98.3%) and ICD codes combined with laboratory values (I^2^= 95.5%). No heterogeneity was detected for algorithms incorporating medication data (I^2^= 0%) (Figure 4).

#### 3.5.2. Systemic Sclerosis

Using ACR/EULAR classification criteria for SSc as the reference standard, meta-analysis was feasible only for algorithms based on a single ICD code. The pooled PPV of a single ICD code was 0.73 (95% CI 0.56-0.84), with substantial between-study heterogeneity (I^2^= 99.3%) (Figure 3C).

## 4. Discussion

This systematic review provides a comprehensive synthesis of studies validating case identification algorithms for CTDs across multiple healthcare data sources. Algorithm performance varied by disease evaluated and algorithm type, with progressively restrictive algorithms consistently improving PPV, although the optimal strategy differed by disease. Meta-analysis of SLE studies demonstrated that algorithms requiring multiple diagnostic codes achieved the highest pooled PPV and best balance between sensitivity and specificity, with the addition of laboratory or medication data not substantially improving diagnostic performance. In contrast, identification of SSc improved with the incorporation of disease-specific features, particularly RP, while for PM/DM, healthcare setting and provider specialty contributed more to diagnostic validity than repeated coding alone. Evidence for SjD was limited to a single study, and no studies were identified for MCTD/UCTD.

This review highlights that optimal case-identification strategies varied according to the clinical characteristics and diagnostic pathways of each CTD, rather than algorithm complexity. The superior performance of repeated diagnostic codes for SLE likely reflects its heterogeneous nature, with multiple healthcare encounters required before diagnosis and the disease’s chronic relapsing nature^58,59^. In contrast, incorporation of RP substantially improved algorithms for SSc, consistent with its role as one of the earliest manifestations of disease and a key predictor of progression^60–63^. For PM/DM, algorithms restricted to specialist or inpatient diagnosis performed best, reflecting the need for specialist evaluation and integration of clinical findings with inpatient investigations such as muscle biopsy^64–66^. Many included studies predated access to myositis-specific autoantibody testing, whose increasing availability has improved disease classification and may enhance future algorithms^66,67^. Overall, these findings suggest that EHRs algorithms should be tailored to the clinical and diagnostic characteristics of individual diseases rather than applying a universal approach or increasing algorithm complexity across CTDs.

The incremental contribution of laboratory and clinical features differed across diseases. In SLE, commonly available laboratory markers such as ANA positivity, provided modest improvement beyond repeated diagnostic codes. This likely reflects the high sensitivity but limited specificity of ANA and the widespread use of serological testing during evaluation of patients with suspected autoimmune diseases, reducing their discriminatory capacity when used alone^68,69^. Similarly, medication records, particularly hydroxychloroquine, may reflect treatment initiated in patients with suspected autoimmune disease or overlap CTDs rather than a confirmed diagnosis of SLE, thereby limiting their discriminatory capacity^70–72^. In contrast, incorporation of RP improved case identification in the available studies. Although RP is not unique to SSc, it represents one of the earliest clinical manifestations of the disease and, when combined with ANA positivity and repeated diagnostic codes, substantially increases the likelihood of true SSc, consistent with the VEDOSS prospective cohort studies of patients with RP^60–62,73^.

Algorithms restricted to inpatient diagnosis consistently demonstrated high specificity and PPV, particularly for SLE and PM/DM, reflecting the fact that hospital-based algorithms preferentially identify patients with severe disease manifestations requiring specialist management, rather than the full spectrum of disease^74–79^. Therefore, although inpatient algorithms provide greater diagnostic certainty, they may be less representative for epidemiological studies aiming to identify the broader disease populations. Consequently, case-identification algorithms should be selected according to the purpose of the study, balancing diagnostic certainty against population representativeness.

An additional methodological consideration relates to the reference standards used across studies. Although many studies relied on ACR/EULAR criteria, these classification criteria were developed to identify homogeneous patient populations for clinical research rather than to establish diagnosis in routine practice^68,80,81^. Consequently, classification criteria may preferentially identify patients with established or more severe disease while underrepresenting individuals with early, incomplete, or atypical presentations^81^. This distinction may influence estimates of diagnostic accuracy, as algorithms validated against classification criteria are likely to perform best for identifying patients meeting research definitions rather than the broader spectrum of disease found in clinical practice^68,80,81^.

Our findings are broadly consistent with previous systematic reviews of rheumatic disease case-identification algorithms. Both Moores et al^10^ and Widdifield et al^15^ reported that requiring repeated diagnostic codes or restricting case identification to specialist/inpatient diagnostic codes generally improved PPV and specificity, whereas increasingly restrictive algorithms often reduced sensitivity. These earlier reviews were based almost exclusively on algorithms using diagnostic codes and highlighted the lack of evidence evaluating the incremental value of laboratory and medication data, as well as the absence of studies using ICD-10 codes^10^. Our review substantially expands this evidence base by including 38 studies across CTDs, incorporating more recent research evaluating ICD-10 code-based algorithms, combinations of diagnostic codes with laboratory, medication, and clinical features, and computational phenotyping approaches. Although evidence for these more complex algorithms remains limited, our findings suggest that laboratory and medication data alone provide only modest improvements over repeated diagnostic codes for SLE, whereas disease-specific clinical features may offer greater accuracy for selected diseases such as SSc.

An important knowledge gap identified by this review is the limited evidence on algorithms for identifying SjD and MCTD/UCTD, with no studies identified for the latter conditions. This may partly reflect the diagnostic and longitudinal complexity of these diseases, which can complicate their identification using EHRs^82–84^.

To our knowledge, this is the first systematic review to comprehensively evaluate and quantitatively synthetise EHR-based algorithm accuracy across multiple CTDs. Includes both ruled code-based algorithms and more recent computational phenotyping approaches, and provides pooled estimates of PPV, sensitivity, and specificity where sufficient data were available. However, several limitations are acknowledged. Considerable methodological heterogeneity was observed across studies, including differences in populations, healthcare settings, coding systems, reference standards, validation methods, and algorithm definitions, which limited direct comparisons. Furthermore, relatively few studies reported sufficient data to assess sensitivity and specificity, as most studies focused on estimating PPV by confirming cases identified through diagnostic codes. This limited meta-analyses of sensitivity and specificity. Evidence for algorithms incorporating laboratory and medication data, clinical features, or computational approaches remained limited across most CTDs. In addition, many studies were conducted in tertiary-care centres or selected clinical populations, which may limit generalisability of the findings to broader healthcare settings.

Future research should prioritise the development and external validation of algorithms in representative healthcare populations, particularly for understudied CTDs such as SjD, MCTD and UCTD. Research ideally should adopt more rigorous methodological approaches, including verification of all or a representative sample of eligible patients, complete reporting of accuracy measures, and standardised descriptions of algorithm components, especially for computational approaches, to facilitate reproducibility and meta-analysis. As healthcare data become increasingly rich, future algorithms should evaluate the incremental value of longitudinal clinical data, disease-specific biomarkers and clinical features, and unstructured EHR data through externally validated computational phenotyping approaches.

## 5. Conclusions

The optimal strategy for identifying CTDs in EHRs differs according to the disease being studied and should be tailored to the clinical and diagnostic characteristics of each disease, and intended research purpose. Repeated diagnostic codes provide a robust approach for SLE, whereas disease-specific clinical features improve case identification for SSc, and specialist or inpatient diagnoses appear particularly valuable for PM/DM. More complex algorithms incorporating laboratory data, medications, or machine-learning approaches may improve accuracy in selected settings; however, current evidence remains limited and requires independent external validation. Future efforts should focus on developing disease-specific and purpose-specific electronic approaches that balance diagnostic accuracy with population representativeness.

## Data Availability

This research is based on published literature. All data used in this research are already included in the article or the supplementary material. The data extraction template is available at the Open Science Framework: OSF | Data Extraction Template.xlsx.

## Acknowledgment

The authors acknowledge the use of large language model to assist with English grammar and spelling during the preparation of the manuscript.

